# Machine learning prediction of motor response after deep brain stimulation in Parkinson’s disease

**DOI:** 10.1101/19006841

**Authors:** J Habets, A Duits, L Sijben, B De Greef, A Mulders, Y Temel, M Kuijf, P Kubben, C Herff, M Janssen

## Abstract

**Introduction:** Despite careful patient selection for subthalamic nucleus deep brain stimulation (STN DBS), some Parkinson’s disease patients show limited improvement of motor disability. Non-conclusive results from previous prediction studies maintain the need for a simple tool for neurologists that reliably predicts postoperative motor response for individual patients. Establishing such a prediction tool facilitates the clinician to improve patient counselling, expectation management, and postoperative patient satisfaction. Predictive machine learning models can be used to generate individual outcome predictions instead of correlating pre- and postoperative variables on a group level.

**Methods:** We developed a machine learning logistic regression prediction model which generates probabilities for experiencing weak motor response one year after surgery. The model analyses preoperative variables and is trained on 90 patients using a ten-fold cross-validation. We intentionally chose to leave out pre-, intra- and postoperative imaging and neurophysiology data, to ensure the usability in clinical practice.

Weak responders (n = 27) were defined as patients who fail to show clinically relevant improvement on Unified Parkinson Disease Rating Scale (UPDRS) II, III or IV.

**Results:** The model predicts weak responders with an average area under the curve of the receiver operating characteristic of 0.88 (standard deviation: 0.14), a true positive rate of 0.85 and a false positive rate of 0.25, and a diagnostic accuracy of 78%. The reported influences of the individual preoperative variables are useful for clinical interpretation of the model, but cannot been interpreted separately regardless of the other variables in the model.

**Conclusion:** The very good diagnostic accuracy of the presented prediction model confirms the utility of machine-learning based motor response prediction one year after STN DBS implantation, based on clinical preoperative variables.

After reproduction and validation in a prospective cohort, this prediction model holds a tremendous potential to be a supportive tool for clinicians during the preoperative counseling.

## Introduction

Subthalamic nucleus deep brain stimulation (STN DBS) is a widely accepted therapy for Parkinson’s disease (PD) patients in which dopaminergic replacement therapy is unsatisfactory.^1-4^ In these patients, DBS can reduce motor symptoms or their fluctuations and thereby improve quality of life.^5^ Despite careful patient selection, some patients still show limited or no improvement of motor disability.^5^ Since the introduction of STN DBS, clinicians aimed to determine reliable predictors.^6^ Researchers have previously studied the predictive power of preoperative clinical information^7^, genetic profiling^8^, MR imaging studies^9^, dopamine-transporter imaging^10^, and intraoperative neurophysiological assessments.^11^

Preoperative levodopa responsiveness of motor symptoms, severity of motor symptoms, and younger age are repeatedly reported as positive predictive factors for postoperative (Movement Disorders Society –) Unified Parkinson’s Disease Rating Scale ((MDS-)UPDRS) motor improvement.^12^ Contrarily, other authors reported preoperative levodopa responsiveness as not being predictive of STN DBS outcome.^7, 13^ Preoperative severe quality of life (QoL) impairment, more time spent in off-condition of dopaminergic medication, levodopa responsiveness, and low BMI have been shown as positive predictive factors on postoperative QoL.^7, 14-16^ Reports on the predictive value of disease duration at surgery, daily levodopa dosage, postural and gait impairment, and non-motor symptoms all show conflicting results.^12, 15, 17, 18^ Comparison of reported motor outcome is hampered due to variance in assessment scales and assessments during varying dopaminergic states.^19^ These non-conclusive results maintain the need for a simple tool which neurologists can use in clinical practice to predict motor outcome after STN DBS for individual patients. To realize a usable and representative tool for the preoperative setting, our approach is limited to preoperative available clinical variables. Prediction during the preoperative phase will always lack information due to surgical factors such as lead placement. This lack of information is inherent to any approach that aims at contributing to a better preoperative counselling.

Machine learning methods are increasingly used in medical practice to unravel patterns and connections to improve prediction in clinical practice.^20^ Predictive machine learning models can be distinguished from traditional statistics by generating outcome predictions for new, individual patients, instead of correlations between pre- and postoperative variables on a group level. The evaluation with cross-validation leads to less limitations in sample size regarding number of considered predictive variables or confounders.^21^ To ensure practical usability, clinical relevance, and interpretable results, the development and implementation of these models requires statistical, programming, and clinical expertise.^22^ Predictive analysis in PD can have additional value by improving challenging clinical decision making instead of only reproducing valid clinical decisions.^23^ One study already applied these methods to predict individual patient response to dopaminergic therapy.^24^ Here, we report the development and proof-of-concept of a prediction model that generates probabilities for weak and strong motor response one year after STN DBS for individual PD patients based on preoperative clinical variables.

## Materials

### Study population

We considered patients who underwent STN DBS for PD in our academic neurosurgical centre between 2004 and March 2018. We included all patients who completed one-year postoperative follow up in this retrospective cohort analysis. We excluded patients who had missing UPDRS-III scores in their preoperative on-medication condition, or postoperative on-medication, on-stimulation condition.

### Surgical procedure

PD patients were indicated for STN DBS based on severe motor symptoms despite optimal levodopa treatment, severe motor fluctuations, or dyskinesia, and often showed a good levodopa responsiveness.^25^ Surgical electrode location was determined based on preoperative MRI trajectory planning, microelectrode recordings, and intra-operative testing. The first part of surgery was performed while the patient was cognizant, after which the lead and pacemaker implantation was completed under general anaesthesia. Postoperative CT examinations verified the electrode location. Postoperative stimulation parameters and dopaminergic drug therapy were set and managed by the neurologist in the outpatient clinic during regular follow-ups.

### Pre- and postoperative variables

All available preoperative demographic data, disease specific data (disease onset, disease duration, levodopa equivalent daily dosage (LEDD))^26^, clinical performance scores ((MDS)-UPDRS, and Hoehn & Yahr (H&Y) scores), as well as relevant neuropsychological assessments in the on-medication condition (Stroop test interference, Verbal Fluency, categorical, and letter tests) were incorporated. We intentionally chose to leave out pre-, intra- and postoperative imaging and neurophysiology data, to ensure the usability in clinical practice. We simulate the situation of a clinician during preoperative counselling. All preoperative clinical and neuropsychological scores were assessed in the on-medication condition except (MDS-)UPDRS III and H&Y scores which were also assessed in the off-medication condition. Preoperative motor levodopa-responsiveness was calculated by subtracting UPDRS III scores in the off-medication condition with scores in the on-medication condition. Postoperative collected variables consist of UPDRS I, II, III and IV and H&Y scores in on-medication and on-stimulation conditions, UPDRS III in on-stimulation and off-medication conditions, and neuropsychological Fluency and Stroop tests in on-stimulation and on-medication conditions. Both MDS-UPDRS and UPDRS scores were collected due to the variation in surgery dates among the population. To create uniform UPDRS scores, MDS-UPDRS scores were recalculated to UPDRS scores.^19^ Pre- and postoperative differences for UPDRS scores I until IV, H&Y scores, LEDD, and neuropsychological assessments were calculated. Furthermore, we registered applied DBS voltage, frequency, and pulse width at one-year follow up. To compare DBS-settings, we computed the mean total electrical energy delivered (TEED).^27^

### Prediction model

We used a machine learning based multivariate logistic regression model that distinguishes itself from (univariate) correlative regression models by predicting individual outcome probabilities (fig. 1). We focused on motor response as outcome and differentiated between ‘strong responders’ and ‘weak responders’. Patients who did not reach a minimal clinically relevant improvement on UPDRS II, III, or IV in on-medication and on-stimulation condition vs. preoperative on-medication condition, were categorized as weak responders. If at least one of the improvements were met, they were categorized as strong responders (fig. 2). The minimal clinically relevant improvement for UPDRS III was 5 points, and for UPDRS II and IV it was 3 points. For further description of this approach, please see Supplementary Material. Our outcome prediction was based on the following available preoperative variables: gender, age at DBS, PD duration at DBS, age at PD onset, UPDRS I, II, III and IV in on-medication condition, motor levodopa response, H&Y scale in on- and off-condition, Stroop interference score, Fluency categorical and total letter score, and the levodopa equivalent daily dosage (LEDD).

**Figure 1.**
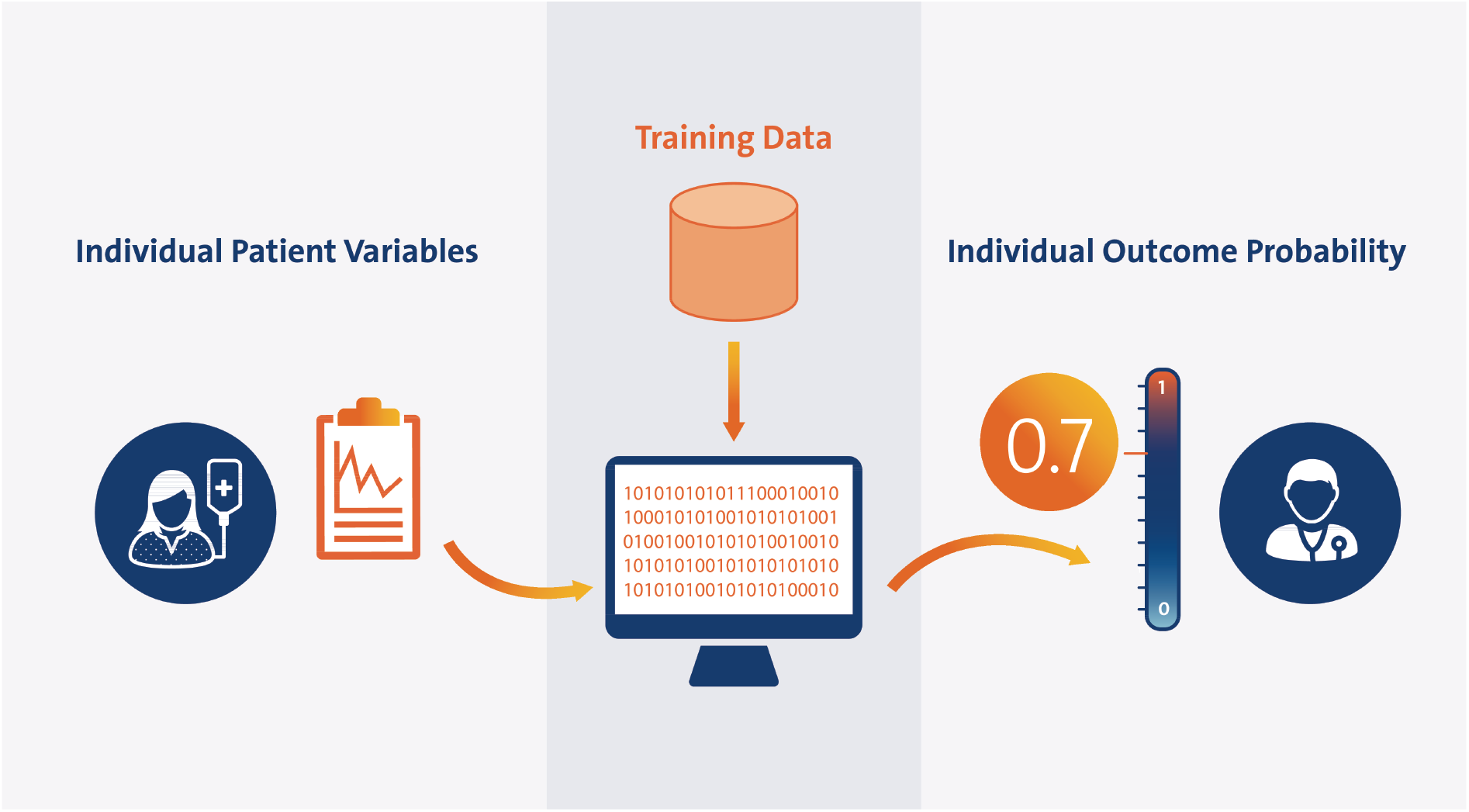
Overview of prediction approach. Workflow of the prediction model as a preoperative counselling tool. The preoperative individual patient variables (left) are inserted in the prediction model, which is trained on the retrospective database (‘Training Data’). The model calculates the probability to become a weak responder between 0 and 1, in this example 0.7. The neurologist can use this probability to inform the patient during preoperative counselling.

**Figure 2.**
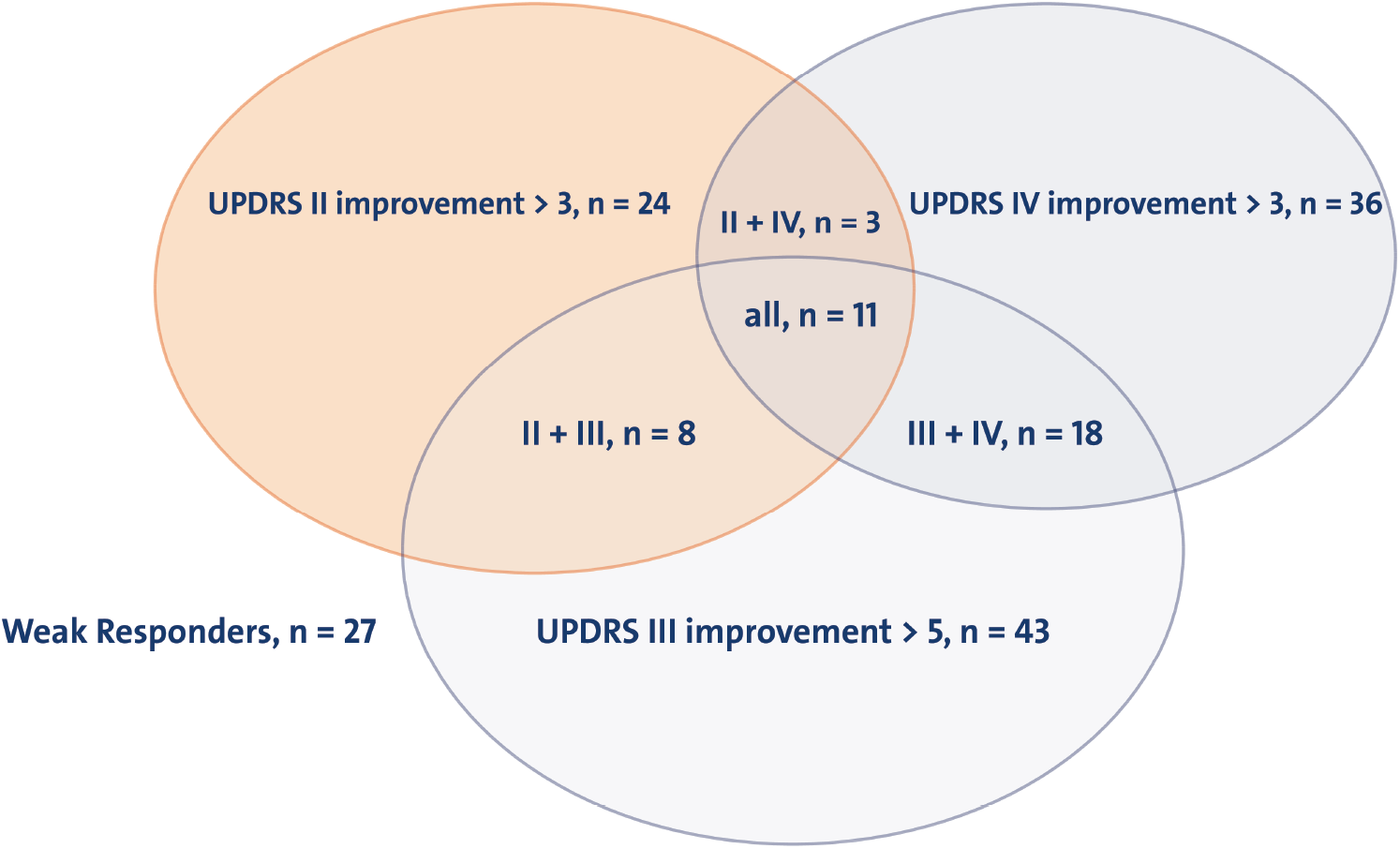
Venn diagram with patient distribution over outcome parameters. The three circles represent the three response criteria, Unified Parkinson’s Disease Rating Scale (UPDRS) I, UPDRS II and UPDRS III, accompanied by the minimal improvement that is considered to be clinically relevant. The numbers within the circles stand for the number of patients that meet the criteria. The overlapping parts of the circles represent the number of patients that meet both or all three criteria.

The predictive logistic regression model was fitted, i.e. trained, on the relation between preoperative variables and postoperative outcome categorization in the retrospective database.^28^ We evaluated our prediction approach in a 10-fold cross-validation. In this approach, the relationship between preoperative variables and postoperative outcome is fitted on 90% of the patients, the ‘training data’. This training assigns a weight, ‘β’, to every single preoperative variable, ‘x’. The prediction model was then evaluated on the remaining 10% of patients in the database, the ‘test data’. When the preoperative variables of a new patient out of the test data were inserted, they got processed with these weights, leading to a prediction of the probability to become a weak responder. The probabilities were generated from the weighted preoperative variables by feeding them into the logistic function 1 / (1 + exp(- β * x)). The fitting and testing procedure was repeated 10 times until every patient was used for testing exactly once. The cross-validation approach simulates validation on new patients. Since the outcome values of the test data subjects were known, the model performance could be tested by comparing predicted probabilities to actual outcome values. We chose logistic regression as a prediction model instead of a deep learning-based model due to the relatively small database size and the fact that the weight, or influence, of every preoperative variable can be interpret separately. for the logistic regression can, which helps to generate an intuition what the prediction is based on.^29^

Since the model predicts probabilities between 0 and 1, the accuracy of the model is strongly dependent on the threshold that decides which probabilities are assumed to be weak responders. To evaluate the overall performance, we employ the receiver operating characteristic (ROC) which visualizes true positive and false positive rates for different thresholds between 0 and 1 (fig 3A). Performance of prediction models is expressed as the area under the curve (AUC) of the ROC (fig. 3A). For converting the model into a prospective application, one of these thresholds should be selected. The model is then expected to produce the true and false positive rates that correspond to the selected threshold.

**Figure 3:**
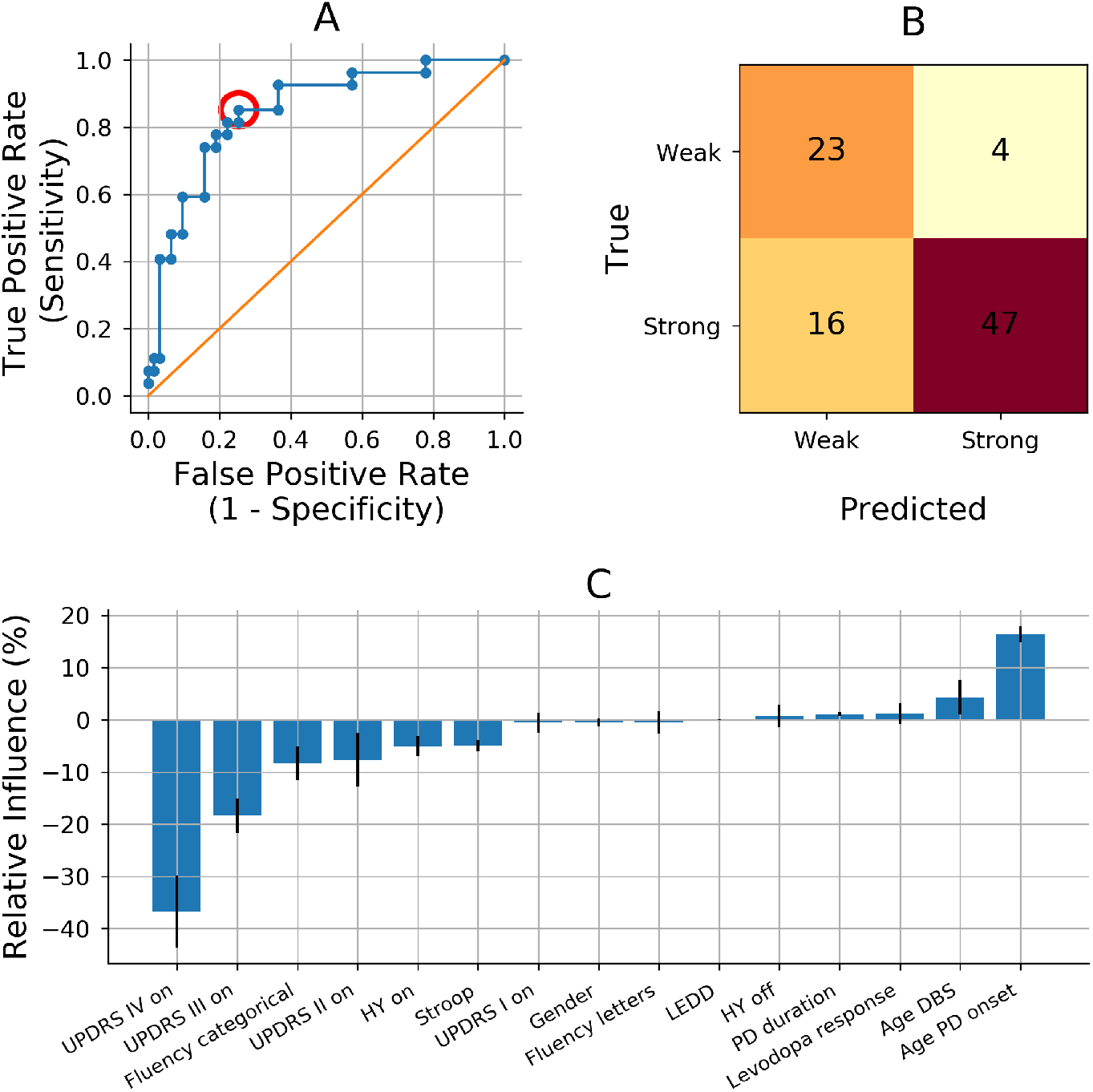
Prediction model performance and importance per predictive variable. A: Visualization of the performance of our prediction model. Our prediction model performs with an average area under the curve (AUC) of the receiver operating curve (ROC, blue line) of 0.88 (standard deviation: 0.14). All the dots on the ROC represent a threshold between 0 and 1 for accepting a probability to be a weak responder to be true. Every threshold leads to a different true positive rate and false positive rate. The red circle represents the threshold corresponding with pane B. The orange line represents chance level in which true positive rates equal true negative rates. B: Matrix of the example when 0.24 is chosen as a threshold for accepting the probability to be a weak responder (red circle in pane A). The true positive rate of 0.85 results in 23 out of 27 true weak responders getting a true weak prediction. The false positive rate of 0.25 results in 16 out of 63 true strong responders getting a false weak prediction. C: Relative influence of all preoperative predictive variables. The normalized Odds Ratios represent the effect of a 1 unit increase in the specific variable, while all other variables stay equal. AUC: area under the curve, DBS: deep brain stimulation, H&Y: Hoehn & Yahr scale, LEDD: levodopa equivalent daily dosage, Levodopa response: difference between UPDRS III off-medication minus UPDRS III on-medication; off: off-medication, on: on-medication, ROC: receiver operate characteristic, TEED: total electrical energy delivered, UPDRS: Unified Parkinson Disease Rating Scale, PD: Parkinson’s disease

To understand which variables the prediction model bases its prediction on, we can explore the importance of every preoperative variable. The model weights can be converted to Odds Ratios by calculating exp(β), which we normalize by subtracting 1 to enable interpretation. We interpret these normalized Odds Ratios as relative influence values in percent. The relative influence values denote the change in probability to be a weak responder when the respective variable increases 1 unit and all other variables stay constant.

Comparative descriptive analysis between preoperative and postoperative variables and between weak and strong responders are performed with Mann-Whitney-U-tests. To facilitate prediction models, we imputed missing data-points in preoperative variables. (For further explanation on the Random Forest imputations applied on preoperative variables, please see Supplementary Material). To prevent imputations of variables that are the target of prediction, we did not impute postoperative variables. Analysis is performed in Python Jupyter Notebook 3 (Jupyter Team, https://jupyter.org, revision fe7c2909) using packages pandas (version 0.24.2), Numpy (version 1.16.4), scikit-learn (version 0.21.2), and Scipy (version 1.3.0). We report our findings according to the TRIPOD Checklist for Prediction Model Development.^30^

## Results

### Preoperative and postoperative variables

We included 98 patients with a well-documented one year follow up after STN DBS of which 90 had no missing data points in UPDRS III score in preoperative on-medication condition and postoperative on-medication and on-stimulation condition. We report descriptive statistics containing the original data (no imputed preoperative data). The total group showed statistically significant postoperative improvements in UPDRS III scores, compared with both preoperative on- and off-medication conditions, and in UPDRS IV scores. We observed a significant decrease in LEDD (table 1). Further, there was a significant deterioration in neuropsychological scores on group level.

**Table 1.** Preoperative and postoperative variables of total population.

| | Baseline characteristics \$ | |
| --- | --- | --- |
| Female sex | 37 (41%) |  |
| Age | 61 (8) |  |
| Disease duration | 10.6 (5.1) |  |
| Preoperative UPDRS III levodopa response | -18.9 (13.4) |  |
| Preoperative UPDRS III % levodopa response | -45.4 (37.9) |  |
| | Preoperative \$ | 1 year follow up \$ |
| UPDRS I ¶ | 1.3 (1.3) | 2.9 (3.1) |
| UPDRS II ¶ | 9.8 (6.5) | 9.7 (5.5) |
| UPDRS III ¶ | 21.8 (12.5) | 16.6 (10.0) * |
| UPDRS III ¶¶ | 39.4 (13.2) | 22.4 (12.4) * |
| UPDRS IV ¶ | 5.5 (4.0) | 2.7 (2.4) * |
| H&Y 1 ¶ | 2 ( 2%)† | - |
| H&Y 1.5 ¶ | 2 ( 2%) | 1 ( 2%) |
| H&Y 2 ¶ | 14 (16%) | 3 ( 5%) |
| H&Y 2.5 ¶ | 34 (40%) | 22 (37%) |
| H&Y 3 ¶ | 24 (16%) | 17 (29%) |
| H&Y 4 ¶ | 9 (10%) | 14 (24%) |
| H&Y 5 ¶ | - | 2 ( 3%) |
| Fluency total categories ¶ | 39.7 (9.4) | 33.7 (9.7) * |
| Fluency total letters ¶ | 35.8 (11.2) | 31.6 (12.0) * |
| Stroop interference ¶ | 56.0 (34.9) | 76.4 (62.8) * |
| LEDD (milligrams) | 1197 (622) | 665 (513) * |
| TEED | - | 134 (130) |

H&Y: Hoehn & Yahr scale, LEDD: levodopa equivalent daily dosage, off-/on-med: off-/on-medication, off-/on-stim: off-/on-stimulation, TEED: total electrical energy delivered, UPDRS: Unified Parkinson Disease Rating Scale
\$. values are given as mean and standard deviation of the mean.
¶: preoperative: on-medication, postoperative: on-medication and on-stimulation
¶¶: preoperative: off-medication, postoperative: off-medication and on-stimulation
†: percentage of Hoehn and Yahr scales are relative based on the number of available data (on: n = 86, off: n = 60)
\*: significant difference with p-value < 0.05, calculated with Mann Whitney-U test

The categorization resulted in 63 strong responders and 27 weak responders (fig. 2). The groups had significant differences on all postoperative UPDRS scores and differences, except for the UPDRS III during on-stimulation and off-medication state (table 2). We observed no significant or relevant differences between the groups regarding neuropsychological scores, LEDD, or TEED.

**Table 2.**
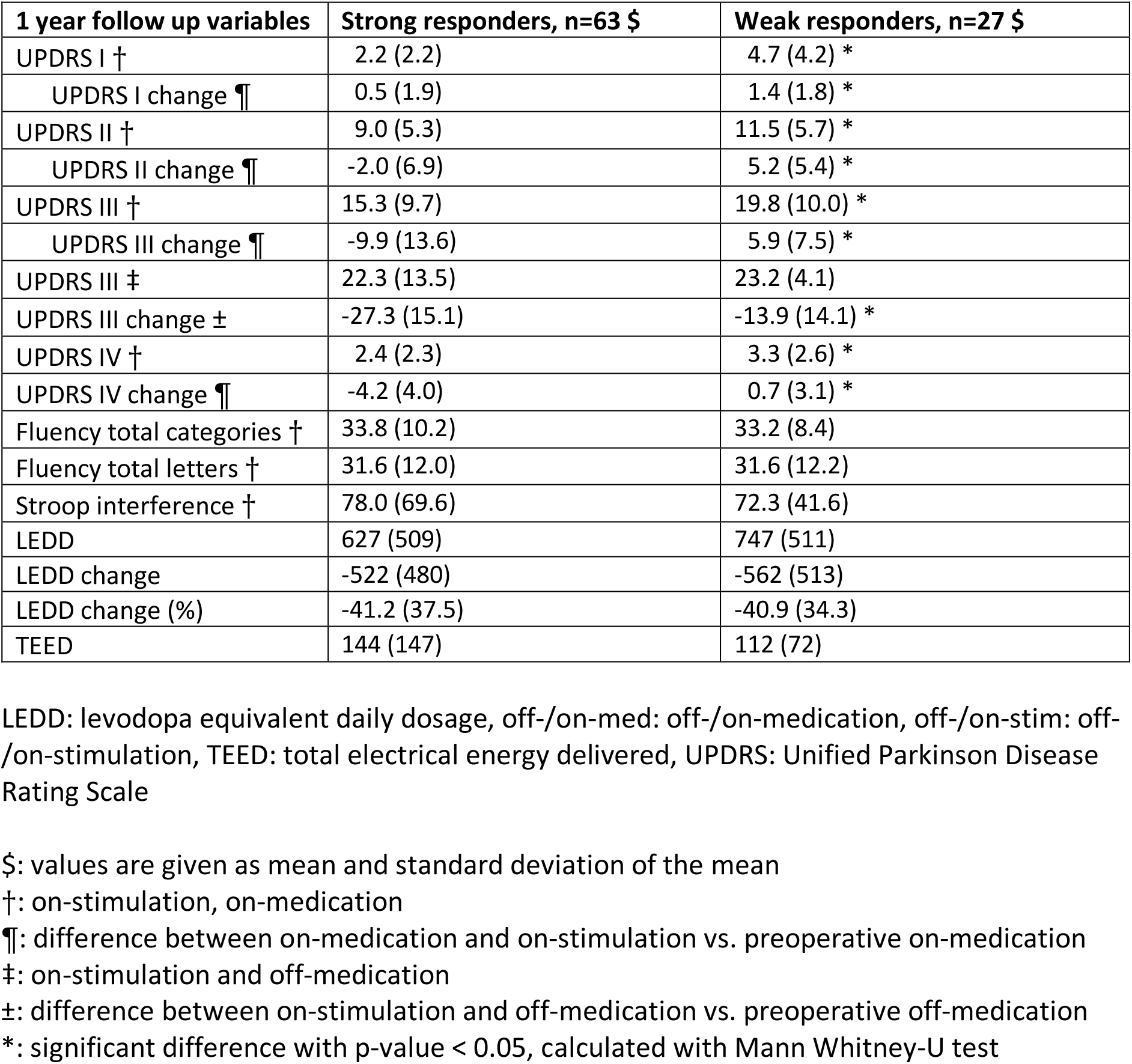
Comparison of postoperative variables in groups with strong responders and weak responders.

### Performance of the prediction model

The receiver operating characteristic (ROC) represents the true positive and false positive rates corresponding to different thresholds that can be chosen to accept a probability to be true. Our prediction model performs with an average AUC of the ROC of 0.88 (standard deviation: 0.14) (figure 3A).

When 0.24 is chosen as a threshold for accepting probabilities to become a weak responder, this leads to a true positive rate of 0.85 and a false positive rate of 0.25 (fig. 3A-B). This also leads to a diagnostic accuracy of 78%, since 70 out of 90 patients are predicted correctly.

The calculated relative influence values represent the importance of each preoperative variable in generating the probability to be a weak responder for each individual (fig. 3C). Older age at PD onset has the strongest relative influence for becoming a weak responder. Similarly, age at DBS is a predictor for becoming a weak responder but is less strong compared to age at PD onset. High preoperative UPDRS III and IV scores in the on-medication condition are the strongest predictors for becoming a strong responder (figure 3C). Additionally, a high preoperative UPDRS II score, high neuropsychological scores on the categorical Fluency test and Stroop test, and higher H&Y score in the on-condition were moderate predictors for becoming a strong responder.

## Discussion

### The clinical value of preoperative prediction of STN DBS motor response

Establishing an accurate prediction tool for motor outcome after STN DBS facilitates the clinician to improve patient counselling, expectation management, postoperative patient satisfaction, and potentially even patient selection.^32^ Due to the complexity and heterogeneity of individual STN DBS candidates, outcome prediction needs to be accompanied by a clinical expert’s appraisal. The likelihoods provided by our approach might also assist clinicians with less experience to identify promising candidates for DBS procedures. Moreover, the accuracy of a prediction model solely regarding clinical preoperative factors will always be limited due to the influence of surgical factors. Nevertheless, we intentionally chose to leave pre-, intra- and postoperative imaging and neurophysiology variables out of our model. This way, we ensure the model to be usable in and representative for clinical practice. We aim to provide the clinician during preoperative counselling with numerical support and insight into the motor outcome which is most likely to expect for an individual patient. This individual estimation can be difficult and has to be made on clinical experience so far.

### The additive value of machine learning methods for clinical prediction models

The applied predictive multivariate logistic regression model was chosen to overcome limitations inherent to conventional (univariate) logistic regression models.^7, 14, 15^ Conventional analyses mainly result in a correlation between one preoperative variable and a postoperative outcome while controlling for several confounding preoperative variables. In designing this analysis, a researcher has to make concessions in selecting the confounding variables because the number of allowed confounding variables is restricted depending on the sample size.^21^ The selection-bias that occurs during this selection directly influences the results and is a major limitation of these conventional statistical models.

The presented model distinguishes itself in this regard, and although it is trained using the same principles of logistic regression, the multivariate approach in our model evaluates all available variables simultaneously. Additionally, by applying cross-validation, we ensure that no overfitting occurs. Moreover, the restriction due to sample size is small and prevents an a priori selection-bias. Lastly, we stress the importance of the use of interpretable predictive machine learning models towards ensuring clinical validity.^29^

### The prediction model performance and its relevant predictive variables

The high AUC of the ROC corresponds to predicted outcome probabilities strongly corresponding to actual outcomes, leading to a very good diagnostic accuracy (AUC = 0.88) (fig. 3A). These results of the presented model confirm the proof-of-concept of machine learning prediction of postoperative motor outcome based on preoperative clinical variables. The prediction accuracy of 78% endorses the potential and need for a further validation in a larger and multicentre patient population. In a future prospective prediction model the nature of the inaccuracy, the balance between false positive and false negative predictions, can be directed by determining the threshold for acceptance (fig. 3B).

The reported influence of higher age at PD onset on becoming a weak responder contradicts earlier findings that report younger age to be a positive predictor for a favourable outcome, although the same meta-analysis reports longer PD duration as a predictor for favourable outcome.^12^

Preoperative UPDRS III and IV scores in the on-medication condition have the highest relative influence values for becoming a strong responder in this model. The finding of high preoperative motor severity increases chances on a stronger response are in line with a meta-analysis, although most included studies report on preoperative severity on off-medication condition.^12^ Evidence on the predictive value of symptom severity in on-medication condition is limited. The finding H&Y scores are not majorly influencing outcome probabilities is in line with literature describing disease severity to positively influence the chance on strong motor response, while axial and balance problems negatively influence this chance.^12^ Furthermore, predictive analysis on QoL outcomes confirm the predictive value of higher UPDRS III scores.^14, 15^ Conversely, recent studies have failed to show positive predictive value of UPDRS III on QoL outcome.^7, 16^

Both high scores on categorical Fluency and Stroop interference are small contributors to becoming a strong responder. Such influences of neuropsychological tests are non-conclusive since a high categorical Fluency score and a high Stroop interference score correspond to better and worse neuropsychological functioning, respectively.

A high UPDRS III levodopa response did not show to influence the probability of becoming a weak responder which contradicts other studies that had identified levodopa responsiveness as an important predictor,^12, 15^ but supports other findings which do not assert predictive value to levodopa responsiveness.^7, 13^

Previous studies have also suggested lower scores on preoperative QoL-scales predict large QoL improvement.^7, 18^ The absence of a predictive value of UPDRS II and III suggests low correlations between UPDRS II & III and the QoL-scale.^7^ We did not reproduce the positive predictive value of less levodopa exposure on QoL improvement.^18^ Regarding levodopa exposure, one should consider that LEDD is expressed in milligrams, which means that the relative influence of a unit increase (1 milligram) is not a clinically relevant increase. Likewise, the absence of a proper non-motor symptom scale hampered potential reproduction of the recently described importance of non-motor symptoms.^17^

The reported influences of the preoperative variables on the outcome probability are partly consistent with the literature, and are partly contradicting literature. We stress that these influences cannot be seen outside the scope of this model. They are only reported to gain insight in the underlying weights which determine the probabilities.

The overview of interpretable weight of each predictive variable is an advantage of the predictive logistic regression model (fig. 3C). This advantage enables clinicians to verify whether the ratio behind the predictions is clinically valid or whether predictions are based on unexpected variables. As stated earlier, the variable weights are not explicitly intended to be interpret outside of the scope of this model. They cannot be interpreted on their own within individual patients when other variables in the model are disregarded.

### The applied categorization for postoperative motor response

To identify the suboptimal responding minority of STN DBS patients objectively, we created a holistic categorization based on available variables representing ADL, motor symptoms, and adverse effects (fig. 2). Previous literature describes STN DBS outcome with a heterogenous variety of variables including UPDRS III severity in on- or off-conditions, time spent in on-condition, and QoL scales.^2, 3, 5, 7^ In the absence of a validated QoL scale, we consider our categorization as the best possible definition of general unsatisfactory outcome.

This categorization resulted in 30% weak responders, which is comparable to reported improvement ratios on quality of life after STN DBS.^5, 14, 17, 18^ Especially since we regarded postoperative differences in on-medication conditions instead of off-medication conditions; and postoperative surgical results are often comparable to the best state in on-medication condition.^12^

The strong and weak responder groups significantly differed on all UPDRS changes, except for UPDRS III scores in the on-stimulation and off-medication condition compared to the preoperative off-medication condition (table 1). A plausible explanation is that nearly all patients will benefit from stimulation compared to the untreated preoperative condition. We consider comparing on-medication states as more natural and realistic because therapy aims to keep patients in on-medication state most of the time.

Given the concordance regarding preoperative and postoperative UPDRS scores and LEDD amounts between our cohort and the literature, we consider our cohort to be representative for the general STN DBS population.^2, 5^ Our cohort showed an expected older age and more severe pre- and postoperative characteristics then a cohort included based on early motor complications.^4^ The observed neuropsychological deteriorations after STN DBS are also in line with previous findings.^31^

## Limitations

Our study is limited by its retrospective character. Missing preoperative data points were overcome by imputations. Outcome values were not imputed to prevent training of the model based on imputed self-generated data. Even though the imputation method was sound, the imputed values will never reach true values and will continue to influence outcomes. Lastly, our cohort lacks QoL outcome variables; our holistic outcome categorization is theoretically a surrogate for QoL but we are unable to make any conclusive claims on QoL.

Second, the accuracy of our prediction approach will always be limited by the fact we only consider preoperative clinical variables. We aim to empower their motor outcome prediction which is currently often made on clinical experience. It is expected that our prediction model cannot perform perfectly, as DBS outcome is also influenced by surgical factors such as lead placement, which cannot be foreseen during preoperative counselling. A prediction in the preoperative phase will therefore always contain variance in accuracy.

## Conclusion

The presented model predicted individual outcome probabilities for becoming a weak responder one-year after STN DBS therapy with very good diagnostic accuracy. This confirms the proof-of-concept of machine learning prediction of motor outcome after STN DBS based on preoperative clinical variables.

The reported influences of the preoperative variables cannot be interpreted outside the scope of this specific prediction model, but endorse the clinical reliability of the applied method.

## Data Availability

Data and analysis scripts are available on reasonable request.

## Acknowledgements

We would like to thank Jackson Boonstra for proofreading the manuscript.

## Notes

### Competing Interest Statement

The authors have declared no competing interest.

### Funding Statement

Yasin Temel and Pieter Kubben received an unrestricted grant from the Weijerhorst foundation.

### Author Declarations

All relevant ethical guidelines have been followed and any necessary IRB and/or ethics committee approvals have been obtained.

Any clinical trials involved have been registered with an ICMJE-approved registry such as ClinicalTrials.gov and the trial ID is included in the manuscript.

## References

1. Limousin P, Pollak P, Benazzouz A, et al. Effect of parkinsonian signs and symptoms of bilateral subthalamic nucleus stimulation. Lancet 1995;345(8942):91–95.

2. Deuschl G, Schade-Brittinger C, Krack P, et al. A randomized trial of deep-brain stimulation for Parkinson’s disease. The New England journal of medicine 2006;355(9):896–908.

3. Odekerken VJ, van Laar T, Staal MJ, et al. Subthalamic nucleus versus globus pallidus bilateral deep brain stimulation for advanced Parkinson’s disease (NSTAPS study): a randomised controlled trial. The Lancet Neurology 2013;12(1):37–44.

4. Schuepbach WM, Rau J, Knudsen K, et al. Neurostimulation for Parkinson’s disease with early motor complications. The New England journal of medicine 2013;368(7):610-622. 5.

5. Williams A, Gill S, Varma T, et al. Deep brain stimulation plus best medical therapy versus best medical therapy alone for advanced Parkinson’s disease (PD SURG trial): a randomised, open-label trial. The Lancet Neurology 2010;9(6):581–591.

6. Pinter MM, Alesch F, Murg M, Helscher RJ, Binder H. Apomorphine test: a predictor for motor responsiveness to deep brain stimulation of the subthalamic nucleus. Journal of neurology 1999;246(10):907–913.

7. Schuepbach WMM, Tonder L, Schnitzler A, et al. Quality of life predicts outcome of deep brain stimulation in early Parkinson disease. Neurology 2019.

8. Weiss D, Herrmann S, Wang L, et al. Alpha-synuclein gene variants may predict neurostimulation outcome. Movement disorders : official journal of the Movement Disorder Society 2016;31(4):601–603.

9. Koirala N, Fleischer V, Glaser M, et al. Frontal Lobe Connectivity and Network Community Characteristics are Associated with the Outcome of Subthalamic Nucleus Deep Brain Stimulation in Patients with Parkinson’s Disease. Brain topography 2018;31(2):311–321.

10. Nakajima A, Shimo Y, Sekimoto S, et al. Dopamine transporter imaging predicts motor responsiveness to levodopa challenge in patients with Parkinson’s disease: A pilot study of DATSCAN for subthalamic deep brain stimulation. J Neurol Sci 2018;385:134-139. 11.

11. Boex C, Tyrand R, Horvath J, et al. What Is the Best Electrophysiologic Marker of the Outcome of Subthalamic Nucleus Stimulation in Parkinson Disease? World Neurosurg 2018;120:e1217–e1224.

12. Kleiner-Fisman G, Herzog J, Fisman DN, et al. Subthalamic nucleus deep brain stimulation: summary and meta-analysis of outcomes. Movement disorders : official journal of the Movement Disorder Society 2006;21 Suppl 14:S290–304.

13. Zaidel A, Bergman H, Ritov Y, Israel Z. Levodopa and subthalamic deep brain stimulation responses are not congruent. Movement disorders : official journal of the Movement Disorder Society 2010;25(14):2379–2386.

14. Daniels C, Krack P, Volkmann J, et al. Is improvement in the quality of life after subthalamic nucleus stimulation in Parkinson’s disease predictable? Movement disorders : official journal of the Movement Disorder Society 2011;26(14):2516–2521.

15. Frizon LA, Hogue O, Achey R, et al. Quality of Life Improvement Following Deep Brain Parkinson’s Disease: Development of a Prognostic Model. Neurosurgery 2018.

16. Abboud H, Genc G, Thompson NR, et al. Predictors of Functional and Quality of Life Outcomes following Deep Brain Stimulation Surgery in Parkinson’s Disease Patients: Disease, Patient, and Surgical Factors. Parkinsons Dis 2017;2017:5609163.

17. Dafsari HS, Weiss L, Silverdale M, et al. Short-term quality of life after subthalamic stimulation depends on non-motor symptoms in Parkinson’s disease. Brain stimulation 2018.

18. Liu FT, Lang LQ, Yang YJ, et al. Predictors to quality of life improvements after subthalamic stimulation in Parkinson’s disease. Acta neurologica Scandinavica 2018.

19. Goetz CG, Tilley BC, Shaftman SR, et al. Movement Disorder Society-sponsored revision of the Unified Parkinson’s Disease Rating Scale (MDS-UPDRS): scale presentation and clinimetric testing results. Movement disorders : official journal of the Movement Disorder Society 2008;23(15):2129–2170.

20. Meyer A, Zverinski D, Pfahringer B, et al. Machine learning for real-time prediction of complications in critical care: a retrospective study. The Lancet Respiratory medicine 2018;6(12):905–914.

21. Wynants L, Bouwmeester W, Moons KG, et al. A simulation study of sample size demonstrated the importance of the number of events per variable to develop prediction models in clustered data. Journal of clinical epidemiology 2015;68(12):1406–1414.

22. Fundamentals of Clinical Data Science. 1 ed: Springer International Publishing, 2019.

23. Cerasa A. Machine learning on Parkinson’s disease? Let’s translate into clinical practice. J Neurosci Methods 2016;266:161–162.

24. Ballarini T, Mueller K, Albrecht F, et al. Regional gray matter changes and age predict individual treatment response in Parkinson’s disease. NeuroImage Clinical 2019;21:101636.

25. Temel Y, Wilbrink P, Duits A, et al. Single electrode and multiple electrode guided electrical stimulation of the subthalamic nucleus in advanced Parkinson’s disease. Neurosurgery 2007;61(5 Suppl 2):346-355; discussion 355-347.

26. Esselink RA, de Bie RM, de Haan RJ, et al. Unilateral pallidotomy versus bilateral subthalamic nucleus stimulation in PD: a randomized trial. Neurology 2004;62(2):201-207.

27. Koss AM, Alterman RL, Tagliati M, Shils JL. Calculating total electrical energy delivered by deep brain stimulation systems. Annals of neurology 2005;58(1):168; author reply 168-169.

28. Pedregosa F, Varoquaux G, Gramfort A, et al. Scikit-learn: Machine learning in Python. Journal of machine learning research 2011;12(Oct):2825–2830.

29. Rudin C. Stop explaining black box machine learning models for high stakes decisions and use interpretable models instead. Nature Machine Intelligence 2019;1(5):206.

30. Collins GS, Reitsma JB, Altman DG, Moons KG. Transparent reporting of a multivariable prediction model for individual prognosis or diagnosis (TRIPOD): the TRIPOD statement. BMC medicine 2015;13(1):1.

31. Okun MS. Deep-brain stimulation for Parkinson’s disease. The New England journal of medicine 2012;367(16):1529–1538.

32. Lin HY, Hasegawa H, Mundil N, Samuel M, Ashkan K. Patients’ Expectations and Satisfaction in Subthalamic Nucleus Deep Brain Stimulation for Parkinson Disease: 6-Year Follow-up. World Neurosurg 2019;121:e654–e660.

